# Computed-tomography estimates of interaural mismatch in insertion depth and scalar location in bilateral cochlear-implant users

**DOI:** 10.1101/2021.02.26.21252533

**Authors:** Matthew J. Goupell, Jack H. Noble, Sandeep A. Phatak, Elizabeth Kolberg, Miranda Cleary, Olga A. Stakhovskaya, Kenneth K. Jensen, Michael Hoa, H. Jeffrey Kim, Joshua G. W. Bernstein

## Abstract

**Hypothesis:** We hypothesized that the bilateral cochlear-implant (BI-CI) users would have a range of interaural insertion-depth mismatch because of different physical placements or characteristics of the arrays, but less than half of electrodes would have less than 75° or 3 mm of interaural insertion-depth mismatch. We also hypothesized that interaural insertion-depth mismatch would be more prevalent nearer the apex, when electrodes were located outside of scala tympani (i.e., possible interaural scalar mismatch), and when the arrays were a mix of pre-curved and straight types.

**Background:** Brainstem neurons in the superior olivary complex are exquisitely sensitive to interaural differences, the cues to sound localization. These binaurally sensitive neurons rely on interaurally place-of-stimulation-matched inputs at the periphery. BI-CI users may have interaural differences in insertion depth and scalar location, causing interaural place-of-stimulation mismatch that impairs binaural abilities.

**Methods:** Insertion depths and scalar locations were calculated from temporal-bone computed-tomography (CT) scans of 107 BI-CI users (27 Advanced Bionics, 62 Cochlear, and 18 Med-El). Each subject had either both pre-curved, both straight, or one of each type of array (mixed).

**Results:** The median interaural insertion-depth mismatch was 23.4° or 1.3 mm. Relatively large interaural insertion-depth mismatch sufficient to disrupt binaural processing occurred for about 15% of electrode pairs [defined as >75° (13.0% of electrode pairs) or >3 mm (19.0% of electrode pairs)]. There was a significant three-way interaction of insertion depth, scalar location, and array type. Interaural insertion-depth mismatch was most prevalent when electrode pairs were more apically located, electrode pairs had interaural scalar mismatch (i.e., one in Scala Tympani, one in Scala Vestibuli), and when the arrays were both pre-curved.

**Conclusion:** Large interaural insertion-depth mismatch can occur in BI-CI users. For new BI-CI users, improved surgical techniques to avoid interaural insertion-depth and scalar mismatch is recommended. For existing BI-CI users with interaural insertion-depth mismatch, interaural alignment of clinical frequency allocation tables by an audiologist might remediate any negative consequences to spatial-hearing abilities.

## I. INTRODUCTION

The purpose of a single cochlear implant (CI) is to partially restore sound and speech perception to people with poor hearing, typically those with bilaterally moderate to profound hearing loss. Recently, bilateral CIs (BI-CIs) have become increasingly prevalent^1,2^. The primary purpose of BI-CIs is to convey cues for auditory spatial perception. Although BI-CI users have better sound localization and speech recognition in the presence of competing sounds compared to unilateral CI users^3-6^, the spatial-hearing benefits that they receive from two ears are far less than those experience by normal-hearing (NH) listeners^7-10^, and they continue to struggle to communicate in noisy environments^11-13^.

In typical auditory systems, binaural sensitivity is computed in brainstem neurons that receive tonotopically symmetric input from the auditory periphery^14,15^. For BI-CIs to convey maximally useful binaural cues, it is imperative to minimize interaural place-of-stimulation mismatch^14,16-20^. This can be partially achieved by matching the insertion depth (i.e., minimizing interaural insertion-depth mismatch) for the two CIs in a BI-CI user. In reality, perfectly matching insertion depths is difficult to achieve in complex CI surgeries due to a high prevalence of relatively shallow insertion depths^21,22^ and scalar translocations^23^. If information about the interaural insertion-depth mismatch were available, the audiologist could potentially realign the ears using asymmetric frequency mappings in the two CIs. In addition, interaural place-of-stimulation mismatch may occur in the scalar location of electrodes (i.e., interaural scalar mismatch). The electrode array is commonly inserted in Scala Tympani (ST) through the round window and is intended to remain entirely in ST^24^, without damaging the basilar membrane and Reissner’s membrane. It is also possible that the apical end of the electrode array can unintentionally puncture one or both membranes, producing electrodes in Scala Media (SM) or Scala Vestibuli (SV)^24-26^. Finally, it is sometimes necessary to insert the array into SM or SV. While interaural insertion-depth mismatch for BI-CI users has been examined in small-N studies using psychophysical^16-20,27,28^ and electrophysiological techniques^18^, the prevalence and characteristics of this mismatch have not been studied at the population level, and interaural scalar mismatch has not been examined at all.

This study used computed-tomography (CT) scans and computer-model rendering^29-31^ to assess the extent to which interaural insertion-depth mismatch exists in a population of 107 BI-CI users. By characterizing the prevalence of interaural insertion-depth and scalar mismatch, and their dependence on array type and tonotopic location, the intent was to provide guidance to surgeons and audiologists in choosing array types and programming strategies to optimize BI-CI hearing outcomes. First, we expected that BI-CI users would have a range of interaural insertion-depth mismatch because of different physical placements of the arrays. We estimate that the population standard deviation in insertion angle is about 75°, based on the summary of the literature provided in Table 1 of Landsberger et al.^21^. If an individual BI-CI users two insertions were selected at random from this distribution, we would expect a mean absolute interaural mismatch of 86°, with 49% of the population having mismatch larger than 75°. However, the cochleae for a given individual are interaurally symmetric. We therefore hypothesized that the amount of interaural insertion-depth mismatch would be smaller than this estimate.

**Table 1:**
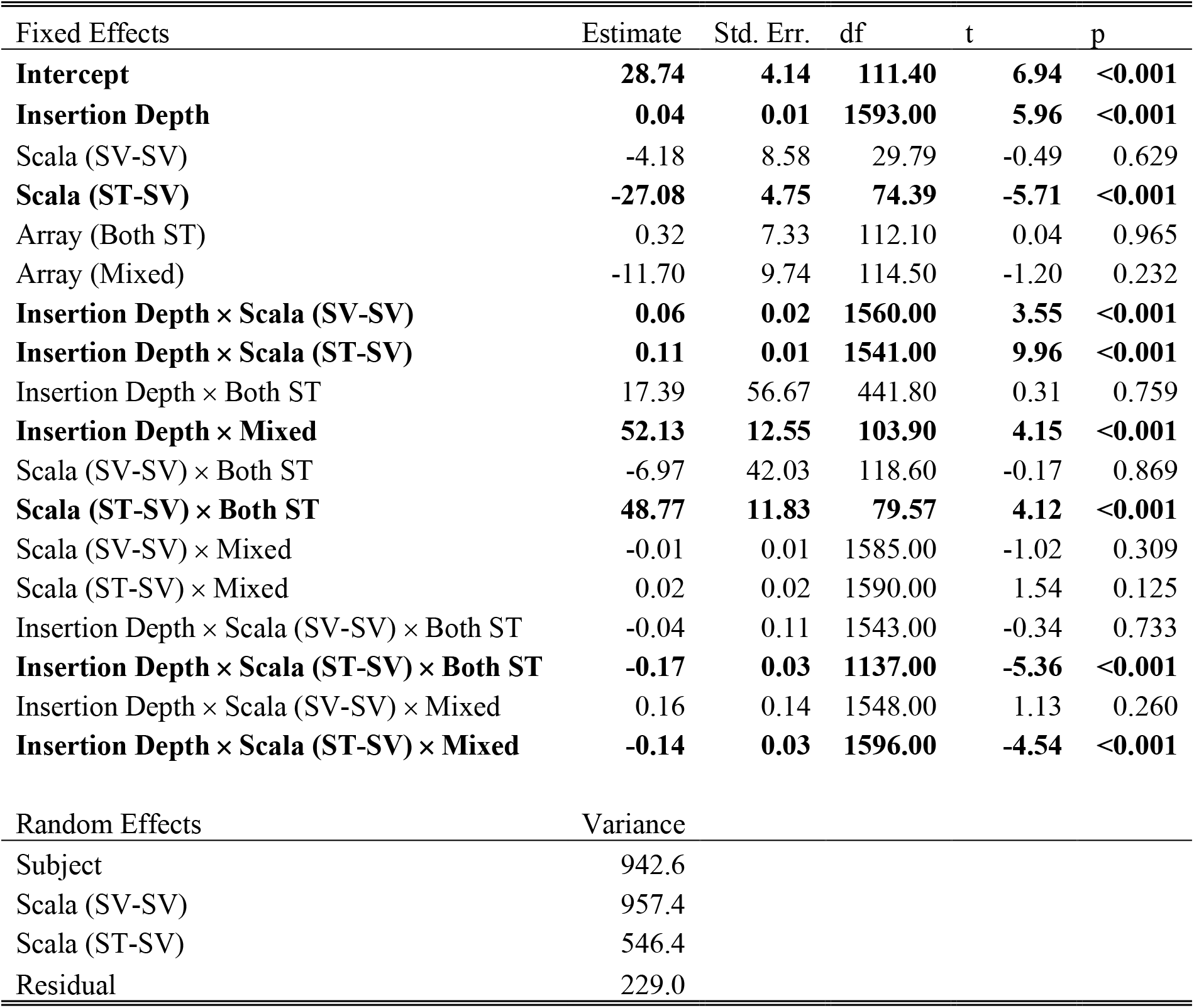
**Results of the linear mixed-effects model. Significant effects and interactions are highlighted in bold**.

Second, we hypothesized that scalar translocation, which has been shown to result in loss of residual hearing^32^ and degradation of speech-in-noise performance^33^, may be prevalent in BI-CI users, particularly at the array’s apical end. This should increase the prevalence of interaural scalar mismatch, particularly in cases when the arrays follow different intended trajectories through the cochlea (i.e., they are a mix of pre-curved and straight types), which should in turn increase the amount of interaural insertion-depth mismatch.

## II. METHODS

Post-operative CT scan and image analysis^29-34^ was performed on 107 BI-CI users from Vanderbilt University and University of Maryland-College Park. The study population included 27 Advanced Bionics users (13 both pre-curved, 10 both straight, 4 mixed), 62 Cochlear Ltd. users (49 both pre-curved, 3 both straight, 10 mixed), and 18 Med-El users (all both straight).

To determine the intra-cochlear location of the electrodes, patient-specific cochlea shape was derived from the patient CT scans using a model that includes segmentation of the intra-cochlear anatomy^29-34^. This analysis quantifies the estimated electrode position in three dimensions. For the present analysis, we were interested in (1) the insertion angle based on a coordinate system defined by a line drawn between the round window and the modiolus and (2) the scalar location of the electrode (ST, SM, or SV)^35^.

Interaural insertion-depth mismatch for number-matched electrode pairs was characterized in three ways. First, insertion depth was estimated in angular degrees around the cochlear spiral, with the modiolus at the center of the coordinate system and the round window defined as zero degrees (horizontal dashed lines in Fig. 1). Second, for comparison with previous estimates^16,17,19,20,36,37^ that binaural processing is tolerant to interaural insertion-depth mismatch <3 mm along an average basilar membrane (BM), insertion angles were translated into BM distances based on the Stakhovskaya^38^ spiral-ganglion correction to the Greenwood^39^ frequency-to-place map, based on a 35-mm cochlear length. For both angular and BM distance, interaural insertion-depth mismatch was calculated as the absolute value of the difference for a given number-matched electrode pair. Third, to provide an acoustic-equivalent measure, interaural mismatch was calculated in octaves based on standard center-frequency (CF) allocation tables for each manufacturer. This interaural mismatch was calculated as the ratio of the CF for a given electrode to the effective CF for an electrode at the same insertion depth in the other ear, obtained by linearly interpolating the electrode insertion angles and allocation tables (log-frequency).

**Figure 1:**
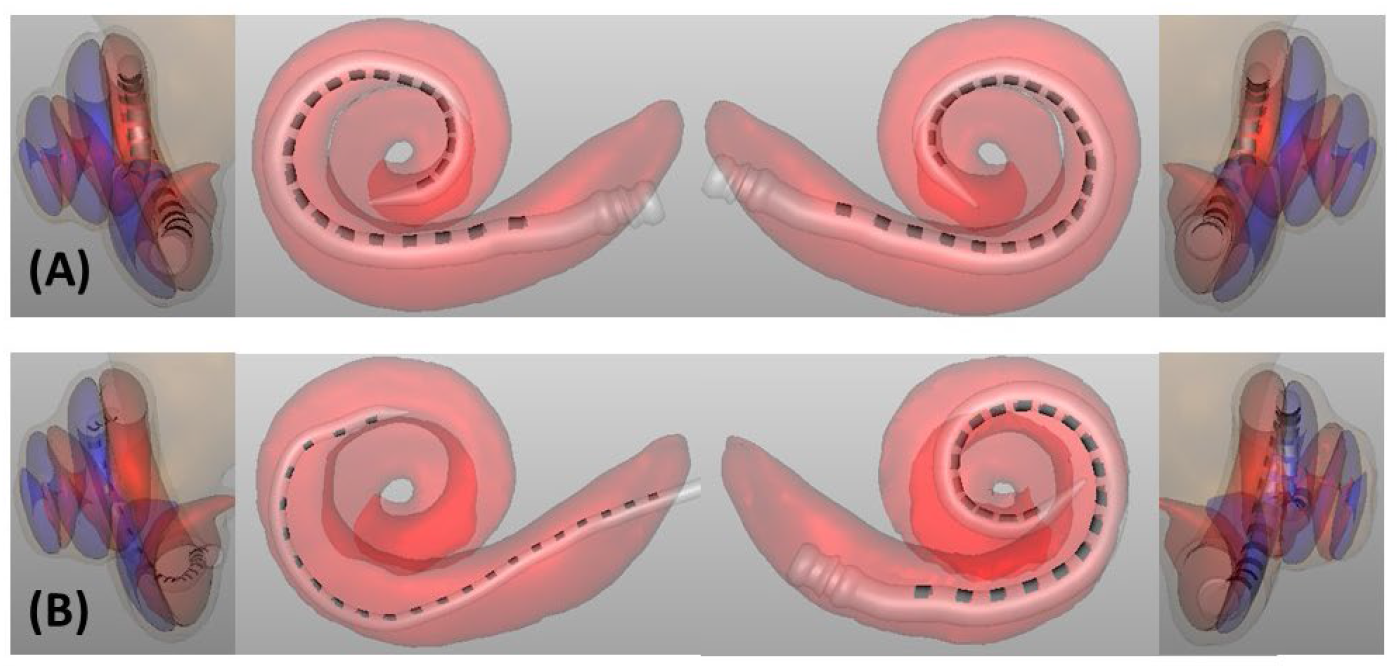
**Two example BI-CI users, with the left two columns showing the left ear and the right two columns showing the right ear. Insertion depth can be seen in the center panels. Scala location can be seen in the leftmost and rightmost panels; ST is shown in red and SV is shown in blue. The example BI-CI user in the top row (A) has a relatively small amount of interaural insertion-depth mismatch (center panels). The electrode arrays are both located in ST (leftmost and rightmost panels), and thus there is no interaural scalar mismatch. The example BI-CI user in the bottom row (B) has a relatively large amount of insertion-depth mismatch (the right-ear array is much deeper than the left-ear array) and there are numerous electrodes located outside of ST (i**.**e**., **some electrode pairs have an interaural scalar mismatch)**.

Electrode scalar location was categorized as ST, SM, or SV based on the anatomical landmarks in the cochlea; ST and SV are shown as different colors in the leftmost and rightmost panels in Fig. 1.

For the analysis in this paper, we focused on ST and SV locations. Electrodes in SM were omitted from the analysis because there were relatively few (7.3%) and because for electrodes near SM, there are a number of different types of trauma that could be occurring, but without direct visualization of scalar structures the specific type is unclear. Some electrodes were also located outside of the cochlea. After removing electrodes pairs where at least one electrode was classified as SM or outside of the cochlea, there were remaining data from 105 subjects and 1766 electrode pairs.

Figure 1 shows CT scans from two example subjects, one with a small amount of interaural insertion-depth mismatch and no interaural scalar mismatch (top row) and one with a large amount of interaural insertion-depth mismatch and an interaural scalar mismatch (bottom row).

## RESULTS

Figure 2 shows probability distribution function (pdf; bars) and cumulative distribution function (cdf; dotted curves) for interaural insertion-depth mismatch expressed in degrees (panel A), mm (panel B), and octaves (panel C) calculated for interpolated electrode pairs with the same electrode number (degrees and mm) or interpolated electrode pairs the same insertion depth (octaves). The median interaural insertion-depth mismatch was 23.4°, 1.3 mm, or 0.27 octaves. Figure 2B includes a vertical line at 3 mm of interaural insertion-depth mismatch, the approximate value where binaural sensitivity statistically degrades in perception studies^16,17,19,20,36,37^. Figure 2A includes a similar vertical line at 75°, which is the average change in insertion angle equivalent to a 3-mm shift along the basilar membrane based on the Stakhovskaya^38^ spiral-ganglion correction to the Greenwood^39^ map. Interaural insertion-depth mismatch >75° occurred for 13.0% of electrode pairs and >3 mm occurred for 19.0% of electrode pairs.

**Figure 2:**
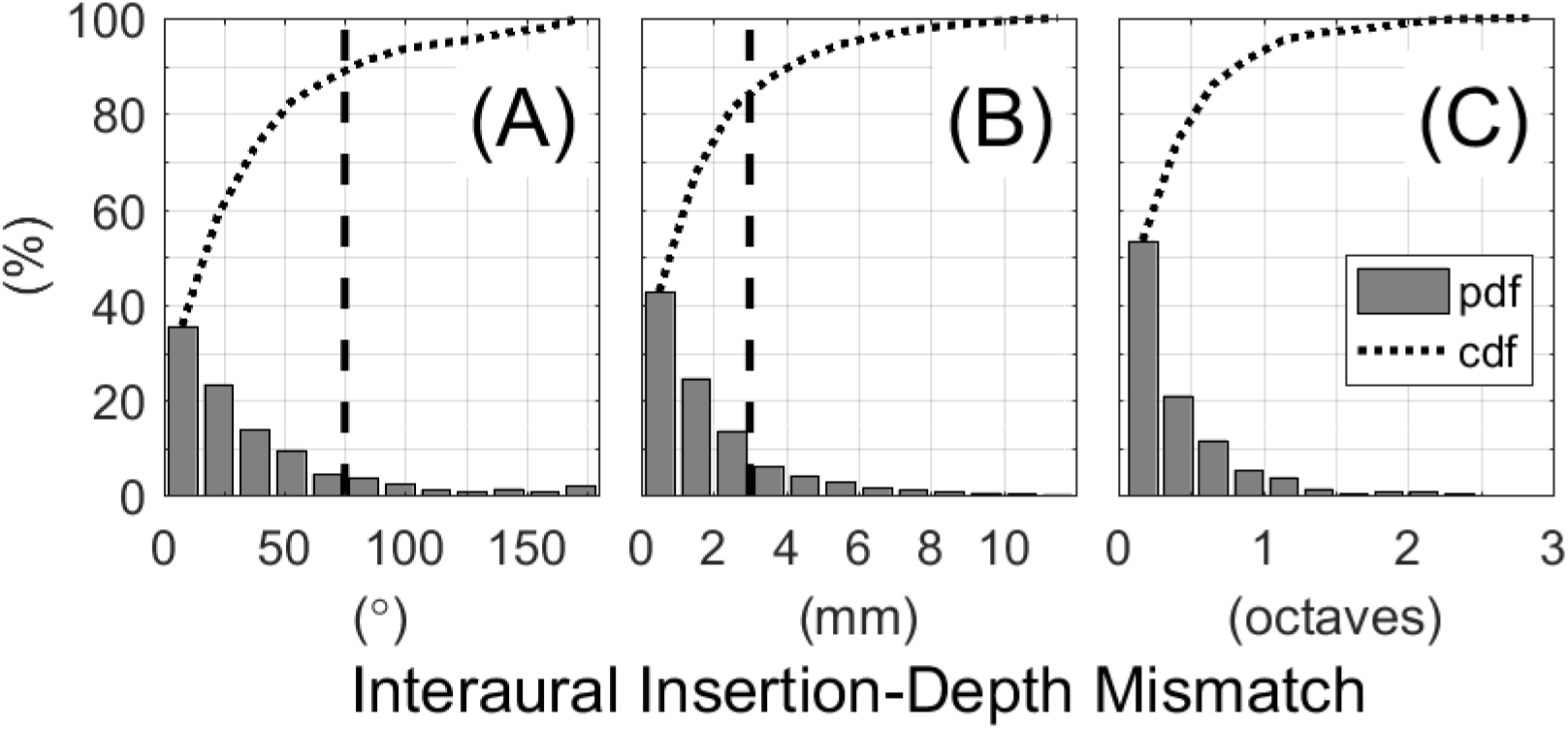
**Distributions of interaural insertion-depth mismatch evaluated with respect to angle (degrees; panel A), distance (mm; panel B), and center frequency (octaves; panel C). Bars represent the probability distribution function (pdf) and the dotted curve shows the cumulative distribution function (cdf; i**.**e**., **the cumulative summation of the pdf). The vertical dashed lines in panels A and B depict an interaural insertion-depth mismatch of 75° and 3 mm, respectively. These represent a relatively large interaural insertion- depth mismatch that would degrade binaural perception as reported in the literature (see text)**.

Figure 3 shows the percentage of individual electrodes located in ST or SV as a function of the insertion depth. There were 2815 electrodes located in ST and 869 electrodes located in SV. The percentage of scalar translocation increased with insertion depth. For depths <180°, fewer than 20% of electrodes were in SV. This increased to 25– 35% for depths between 240°–420° and about 50% for depths >420°.

**Figure 3:**
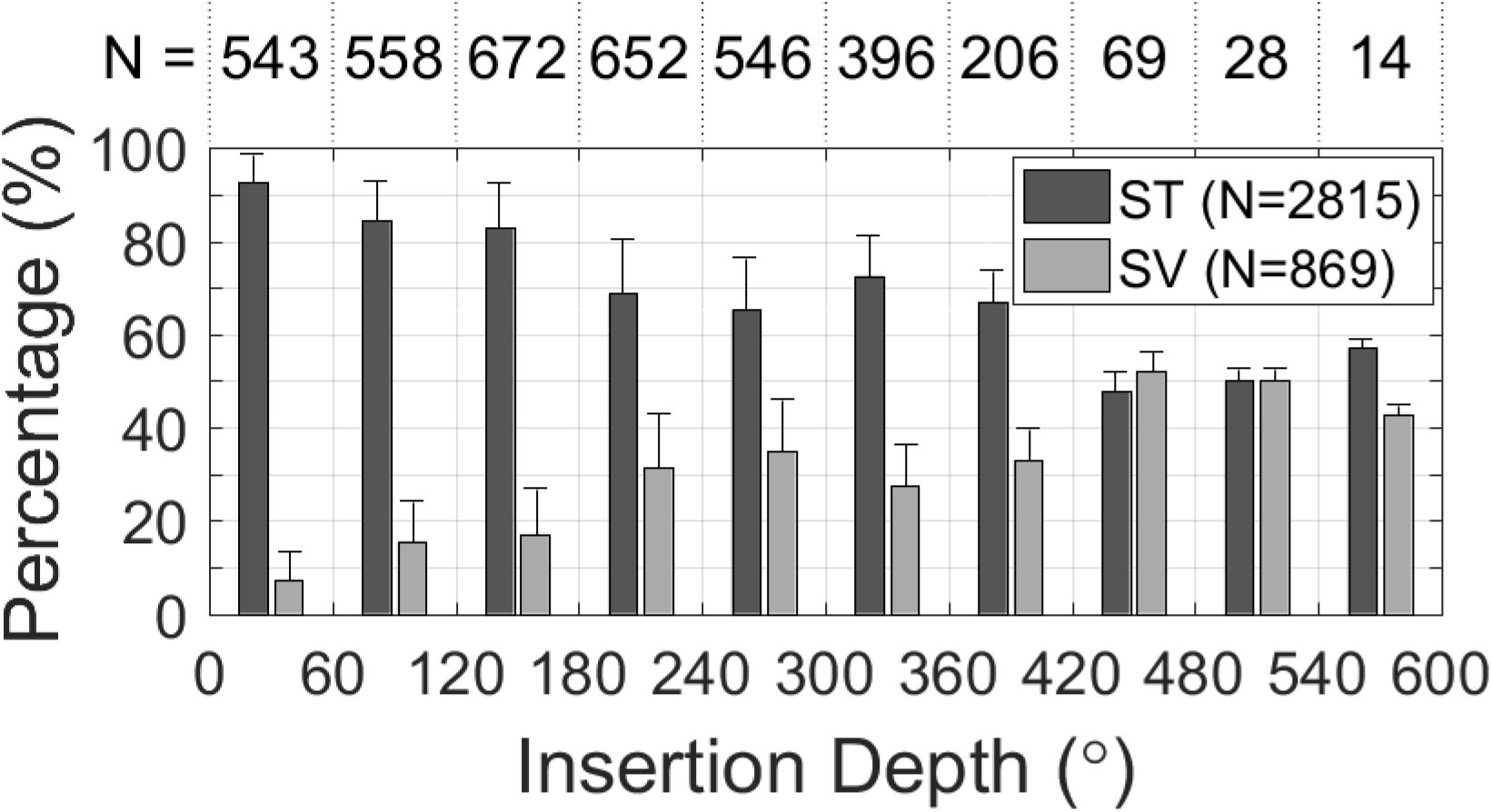
**Histogram of individual electrodes located in Scala Tympani (ST) and Scala Vestibuli (SV) as a function of insertion depth in degrees. The numbers (N) at the top of the plot show the number of electrodes at each depth, and the numbers inside the legend indicate the total number of electrodes in each category. Error bars show +1 standard deviation**.

Figure 4 shows the absolute interaural insertion-depth mismatch as a function of the average insertion depth for each electrode pair, plotted separately for each combination of scalar location (rows) and array type (columns). Figures 4E and 4F had insufficient counts to assess the relationship between interaural insertion-depth mismatch and insertion depth, with only 1 or 2 subjects in each case. For the other categories, the fitted regression lines show a tendency for more interaural insertion-depth mismatch with greater insertion depth (i.e., positive slopes) when at least one of the arrays was pre-curved (left and right columns).

**Figure 4:**
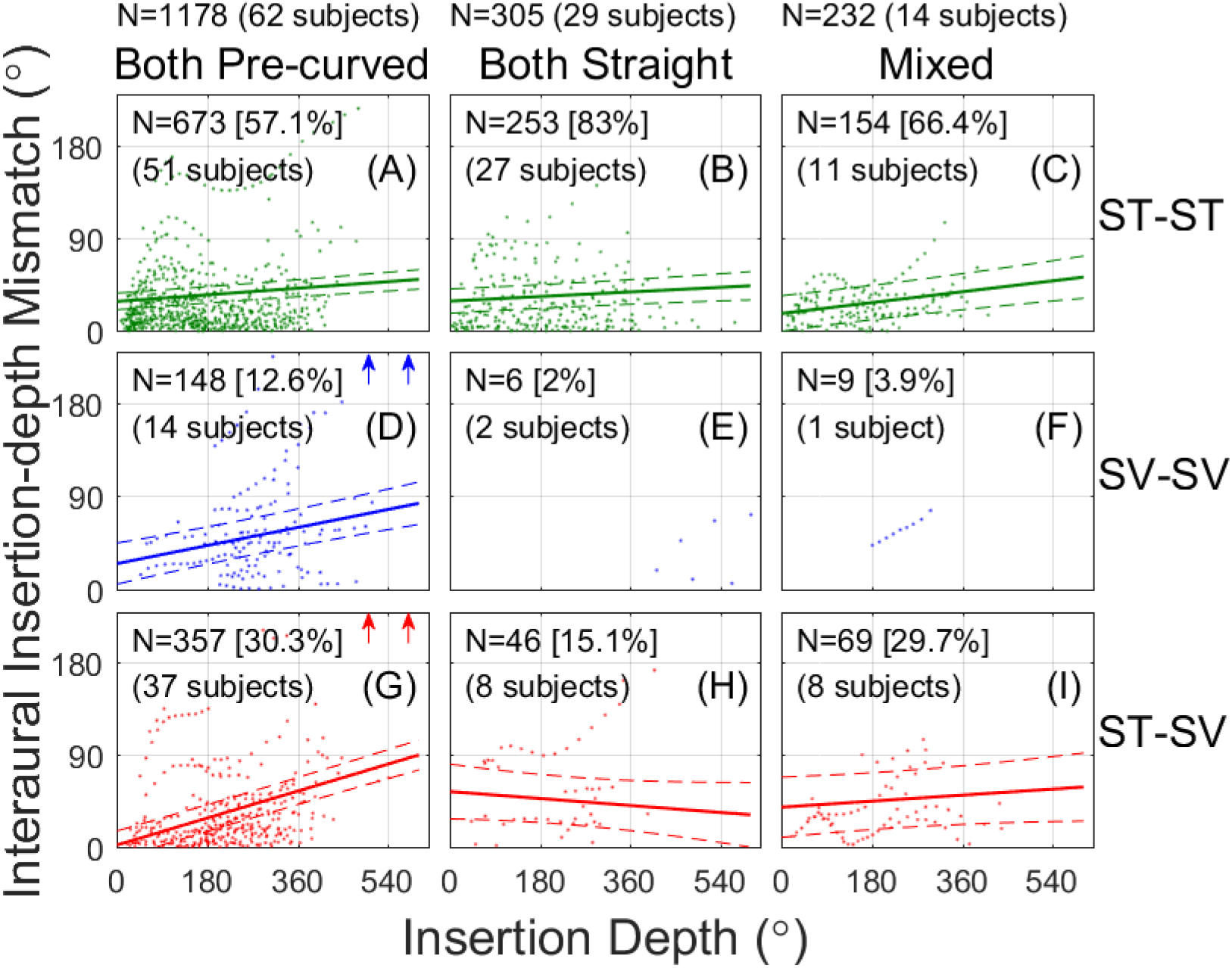
**Interaural insertion-depth mismatch as a function of insertion depth for the different categories of scalar location and array type. Solid lines show linear fits to the data from the linear mixed effects model. Dashed lines show the 95% confidence interval of the fits. Number of electrode pairs (the data points) and number of subjects contributing in each condition is reported in each panel. Arrows indicate conditions where the points with >240° of interaural insertion-depth mismatch occur, but were outside the plot range to better show the fits**.

These trends were analyzed with a linear mixed-effects (LME) model (R version 4.0.3) using the buildmer (v1.7.1)^40^ and lme4 (v1.1-26)^41^ packages. The best-fitting model included interaural insertion-depth mismatch as the dependent variable and three fixed effects: insertion depth (continuous variable), scalar location (categorical variable: ST-ST, SV-SV, and ST-SV), and array type (categorical variable: both pre-curved, both straight, and mixed). Subject and scalar location were significant random effects. For the categorical variables, the model was referenced to the ST-ST and both pre-curved categories.

The results of the main LME analysis are reported in Table I. There were significant main effects of insertion depth on interaural insertion-depth mismatch (more mismatch for more apically located electrodes) and scalar location (cases of interaural scalar mismatch ST-SV showed more mismatch than cases with no interaural scalar mismatch ST-ST). There were, however, also many significant two- and three-way interactions, suggesting that the relationship between interaural insertion-depth mismatch and insertion depth varied across array and scalar categories.

To examine the three-way interactions, seven post-hoc models were run (the previous analysis and six additional models with different reference levels), one for each combination of scalar location and array type, excluding the two SV-SV combinations with insufficient counts/subjects. Bonferroni corrections were applied for seven analyses (criterion: *p*<0.0071=0.05/7). Table II reports the slopes for the different scalar and array combinations. It also reports the difference in slopes of the fits, which was obtained by comparing the interactions between insertion depth and different categorical conditions of the releveled models.

**Table 2:** **Slopes for the different scalar and array categories (left columns) and a comparison of slopes across categories (right columns). Bolded *p* values are significant at the *p*<0.0071 level. PC=pre-curved; ST=Scala Tympani; STR=straight; SV=Scala Vestibuli**.

|  |  |  |  |  | Slope Comparison |  |  |  |  |  |  |  |  |
| --- | --- | --- | --- | --- | --- | --- | --- | --- | --- | --- | --- | --- | --- |
| Slopes |  |  |  |  | ST-ST | ST-ST | ST-ST | SV-SV | SV-SV | SV-SV | ST-SV | ST-SV | ST-SV |
| Scala | Array | Intercept | Slope | $p$ | Both PC | Both STR | Mixed | Both PC | Both STR | Mixed | Both PC | Both STR | Mixed |
| ST-ST | Both PC | 28.740 | 0.036 | **<0.0001** | -- | 0.309 | 0.125 | **0.0004** | -- | -- | **<0.0001** | **<0.0001** | **<0.0001** |
| ST-ST | Both STR | 29.050 | 0.025 | 0.005 |  | -- | 0.038 | 0.733 | -- | -- | **<0.0001** | 0.040 | 0.374 |
| ST-ST | Mixed | 17.032 | 0.059 | **<0.0001** |  |  | -- | 0.260 | -- | -- | **<0.0001** | 0.374 | 0.356 |
| SV-SV | Both PC | 24.554 | 0.098 | **<0.0001** |  |  |  | -- | -- | -- | 0.009 | 0.205 | 0.038 |
| SV-SV | Both STR | -- | -- | -- |  |  |  |  | -- | -- | -- | -- | -- |
| SV-SV | Mixed | -- | -- | -- |  |  |  |  |  | -- | -- | -- | -- |
| ST-SV | Both PC | 1.653 | 0.147 | **<0.0001** |  |  |  |  |  |  | -- | **<0.0001** | **<0.0001** |
| ST-SV | Both STR | 54.105 | -0.038 | 0.194 |  |  |  |  |  |  |  | -- | 0.061 |
| ST-SV | Mixed | 38.721 | 0.033 | 0.173 |  |  |  |  |  |  |  |  | -- |

Figure 4 and Table II show that interaural insertion-depth mismatch increased with increasing insertion depth for four conditions: all three cases with both-pre-curved cases (Fig. 4A, D, G), and the mixed ST-ST case (Fig. 4C). There was no effect of insertion depth in both-straight ST-ST condition (Fig. 4B). In this case, the slope of the regression line was shallow (0.025), meaning that the interaural insertion-depth mismatch increased by only 9° for a full 360° turn around the cochlea towards the apex. There was also no effect of insertion depth in the both-straight ST-SV (Fig. 4H) and mixed ST-SV cases (Fig. 4I), although the confidence intervals for these cases were large because of relatively few data points.

Comparing the slopes (right half of Table II) across the top row of Fig. 4 (ST-ST electrodes) shows no significant effect of array type, while comparing across the bottom row (ST-SV electrodes) shows significantly greater slopes for the both-pre-curved case than for the both-straight or mixed cases. Comparing the slopes down the left column of Fig. 4 (both pre-curved) shows that the slopes were significantly greater for the SV-SV (Fig. 4D) and ST-SV (Fig. 4G) cases compared to the ST-ST case (Fig. 4A). There was a significantly greater slope for the both-pre-curved ST-SV condition (Fig. 4G) than all the other conditions, except for both-pre-curved SV-SV condition (Fig. 4D). There were no significant changes in slope as a function of scalar type down the middle (both-straight) and right columns (mixed arrays) of Fig. 4, although this might be due, at least in part to the relatively few counts for the both-straight ST-SV (Fig. 4H) and mixed ST-SV (Fig. 4I) cases.

To summarize, the significant three-way interaction reflected different dependencies of interaural insertion-depth mismatch on insertion depth. When both electrodes were in ST, there was a slight dependence of mismatch on insertion depth for all array-type categories (Fig. 4, top row). But when both arrays were pre-curved, there was a greater dependence of mismatch on insertion depth when at least one of the electrodes was outside of ST (Fig. 4, left column). The largest dependence on insertion depth occurred for the cases with two pre-curved arrays and an interaural scalar mismatch (ST-SV scalar locations; Fig. 4G). For these cases, the slope was 0.147, which means that the interaural insertion-depth mismatch increased by 52.9° for a full 360° turn around the cochlea towards the apex.

## IV. GENERAL DISCUSSION

### A. Summary of Results and Relationship to Previous Studies

Many factors contribute to relatively small binaural benefits in BI-CI users, including device-related and surgical factors^42-45^. Interaural place-of-stimulation mismatch can reduce binaural functioning in BI-CI users because the binaural differences are computed at the level of the brainstem using frequency-matched inputs^14^. Previously, the prevalence of interaural place-of-stimulation mismatch was unknown as it has never been systematically analyzed in a large number of patients; most psychophysical or electrophysiological studies investigating interaural place-of-stimulation mismatch have involved relatively small sample sizes (N≈10)^17-20,27,28,43,46-54^. Using a larger database of CT scans from N=107 BI-CI users, interaural place-of-stimulation mismatch (which likely is a combination of non-neural and neural factors) was estimated based on non-neural physical interaural insertion-depth mismatch (insertion depth differences of individual electrode pairs, measured in degrees).

Our main finding was that interaural insertion-depth mismatch was fairly common (Fig. 2), where the median interaural insertion-depth mismatch was 23.4° or 1.3 mm. From previous reports, we estimated that less half of electrode pairs would exhibit a relatively “large” amount of mismatch of 75° or 3 mm, the latter quantity has been shown to incur significant decrements in perceptual binaural sensitivity and fusion^16,17,19,20,36,37,55^. We found this to be true, where 13% of electrode pairs had >75° and 19% of electrode pairs had >3 mm of interaural insertion-depth mismatch. It is important to note that the large-amount-of-mismatch value is derived from studies involving single-electrode stimulation in each ear; it has also been suggested that BI-CI users may be less tolerant to interaural insertion-depth mismatch using multi-electrode stimulation^54^. Indeed, it is unclear what to judge as a disruptive amount of interaural insertion-depth mismatch without paired perceptual measurements.

Our second main finding was a significant three-way interaction that revealed larger interaural insertion-depth mismatch for increasing insertion depths, when there was at least one pre-curved array (i.e., both pre-curved or mixed arrays) but particularly when there were two pre-curved arrays, and when the electrodes had an interaural scalar mismatch (Fig. 4, Tables I and II). While this broadly confirms the second hypothesis of the study, our initial hypothesis did not include the both-pre-curved condition, and hence this finding was notable. Consistent with previous reports^24^, the probability of non-ST electrode location for a single array increased with the insertion depth in the current data set (Fig. 3). Pre-curved arrays also tended to have more mismatch (Fig. 4, Tables I and II). When there were two pre-curved arrays and translocation occurs (typically at the apex), this was the instance that caused the most interaural insertion-depth mismatch.

These results might account for some of the dependence of binaural sensitivity on tonotopic place that is sometimes reported in the literature^6^, in particular the tendency for the poorest ITD discrimination performance to occur near the apical end of the array^43,44,56,57^. This may be a result of sensitivity being degraded by translocations^58^, and interaural insertion-depth or scalar mismatch (Figs. 3 and 4), all of which become more prevalent toward the apex. These occurrences might be inconsistent between studies with small study populations, explaining the mixed results in the literature.

### B. Limitations and Future Directions

While we have shown interaural insertion-depth mismatch occurs readily in BI-CI users, this analysis reflects only the anatomical location and does not account for other factors that might affect binaural sensitivity. Peripheral biological factors such as neural degeneration, dead regions, and tissue scarring may alter the relative populations of peripheral neurons being stimulated^59-61^. Furthermore, it is possible that binaural circuits could adapt to address interaural insertion-depth mismatch^62-64^, which could affect binaural functioning and/or binaural fusion^55,65^; however, little physiological evidence has supported such an idea to date^18^.

To more directly assess the relationship between physical and functional mismatch, future work should consider relating binaural sensitivity to CT measurements of intracochlear electrode location. Recent measurements in a smaller group of 20 BI-CI listeners and 23 SSD-CI listeners suggests that CT and ITD-based estimates of interaural insertion-depth mismatch are closely aligned^66^. In contrast, perceptual^18,65,66^ or electrophysiological binaural sensitivity showed incongruence with pitch^18^. Another limitation was that while it included >100 BI-CI users, a larger sample would have provided more statistical power to examine some of the subgroups in this study (e.g., use of two straight arrays, or a mix of pre-curved and straight arrays, Fig. 4).

### C. Clinical Implications

Interaural place-of-stimulation matching has the potential to improve binaural performance in BI-CI users. During implantation, improved surgical techniques to avoid interaural insertion-depth and scalar mismatch are recommended, for example, by using CT-based surgical guidance^67^. Post-implantation, knowing the interaural cochlear locations that maximize binaural sensitivity could guide the audiological frequency-mapping process to optimize spatial hearing. While this might be facilitated by psychophysical and electrophysiological measurements of binaural performance^16-18,27,28,55^, these measurements require ITD sensitivity, which is not possible for some CI users^68-70^, and can be laboriously long^28^. Use of CT scans, on the other hand, are rapid and precise. This approach acknowledges that the binaural hearing system requires coordination across the ears, rather than treating the two devices as separate systems.

When considering bilateral implantation, there are competing issues involved in the selection of an array type. On one hand, some studies suggest that a pre-curved array might provide an advantage for monaural speech-understanding outcomes and provide better channel independence^71,72^, although translocations with such arrays appear to be a potential problem because of decreased performance^58^. On the other hand, our results show that pre-curved arrays are also more likely to result in more interaural insertion-depth and scalar mismatch toward the apex, which might reduce binaural sensitivity, although frequency- mapping programming techniques might address this issue. In any case, it is important to qualify this interpretation because the observed significant differences between array types was small and it is unclear if these differences would substantially affect perception.

## Data Availability

The authors confirm that the data supporting the findings of this study are available within the article.

## ACKNOWLEDGMENTS

We thank Danielle King, Kelly Miller, and Kristina Milvae for help with data analysis. Research reported in this publication was supported by the National Institute On Deafness And Other Communication Disorders of the National Institutes of Health under Award Number R01DC015798 (M.J.G. and J.G.B.W.), R01DC014037 (J.N.), and R01DC014462 (Dawant). The content is solely the responsibility of the authors and does not necessarily represent the official views of the National Institutes of Health. Portions of this work were presented at the 176th Meeting of the Acoustical Society of America, 42nd Midwinter Meeting of the Association for Research in Otolaryngology, and the 19th Conference on Implantable Auditory Prostheses.

The identification of specific products or scientific instrumentation is considered an integral part of the scientific endeavor and does not constitute endorsement or implied endorsement on the part of the authors, Department of Defense, or any component agency. The views expressed in this article are those of the authors and do not reflect the official policy of the Department of Army/Navy/Air Force, Department of Defense, or U.S. Government.

